# Efficacy of Mobile Application Delivered Lifestyle Interventions in Managing Gestational Weight Gain: A Systematic Review and Meta-Analysis with Meta-Regression

**DOI:** 10.64898/2026.05.29.26354025

**Authors:** Gresita Novelia Uirianto, Samuel Partogi Nababan

**Author notes:** Corresponding author: Gresita Novelia Uirianto.

## Abstract

**Introduction:** Managing gestational weight gain (GWG) is crucial for the health of mothers and their children. Mobile applications (apps) specifically designed for pregnancy are emerging as modalities to deliver accessible lifestyle intervention at a low-cost. However, current studies are varied in results and suffer from heterogeneity. Thus, we conducted this systematic review and meta-analysis to summarize the efficacy of mobile apps in managing GWG and investigate variables that may contribute to heterogeneity.

**Methodology:** Seven databases were systematically searched up to 9 November, 2024. Only randomized controlled trials (RCTs) were included. Outcomes were excessive GWG and inadequate GWG according to the 2009 Institute of Medicine (IOM) guideline. Quality appraisal was performed using the Cochrane Risk of Bias 2 (RoB 2) tool. Random-effect model meta-analysis was conducted using odds ratio (OR) as the summary measure alongside their 95% confidence intervals (CI).

**Results and Discussion:** Fifteen RCTs were included. Mobile apps led to a significant overall decrease in excessive GWG (OR: 0.71; 95% CI: 0.54 to 0.95; p-value: 0.02; I^2^: 60%). Subgroup analysis showed that social media apps, self-monitoring functionalities, and overweight/obese patients are associated with a significant reduction in excessive GWG. However, there was significant evidence of small-study bias in the analysis. Moreover, mobile apps also significantly increased inadequate GWG (OR: 1.51; 95% CI: 1.04 to 2.21; I^2^: 0%). Meta-regression did not reveal any significant finding.

**Conclusion:** In conclusion, mobile app interventions are shown to be effective in preventing excessive GWG, particularly social media apps and those with self-monitoring functionalities. However, the reduction in excessive GWG may only be seen in overweight and obese patients and more studies are needed to ascertain this finding. Lastly, mobile apps are associated with an increased risk of inadequate GWG and strategies to combat inadequate GWG are needed.

## INTRODUCTION

Managing gestational weight gain (GWG) is crucial for the health of mothers and their children. Adequate GWG supports fetal development and reduces complications during pregnancy, but gaining too much or too little weight can lead to significant problems. Excessive GWG is linked to conditions like gestational diabetes, high blood pressure, and long-term postpartum obesity for the mother, as well as larger-than-average birth weight and increased obesity risks for the child.^1,2^ On the other hand, inadequate GWG can result in small for gestational age and low birth weights, highlighting the need for effective strategies to help pregnant women maintain a healthy weight.^3^

With the growing use of mobile technology, healthcare delivery has seen a shift toward innovative digital solutions. Mobile applications (apps) specifically designed for pregnancy are emerging as valuable tools to encourage healthy behaviors like proper nutrition and regular physical activity–which are essential for managing GWG–at a high intervention dose and low-cost. These apps offer features such as self-monitoring tools, personalized tips, and educational resources, making them convenient and accessible.^4^ These modalities of mobile apps integrated into pregnancy care could provide distinct advantages, such as improved accessibility and increased user engagement.^5^

Mobile apps have been researched heavily in primary studies. However, the results and characteristics of these mobile app interventions have been varied. Additionally, a previous meta-analysis by Chan et al. suffered from small samples of studies and lack of investigation into the heterogeneity of the intervention characteristics, participant characteristics and study characteristics.^6^ As a result, these meta-analyses suffer from a lack of generalizability and certainty of evidence. Further secondary studies resolving these limitations are needed to deepen the understanding of the effectiveness of mobile apps in managing GWG, especially in investigating what characteristics makes the intervention effective and what other variables can affect it. Thus, this study was conducted aiming to resolve these knowledge gaps.

This study analyzes the effectiveness of mobile app-based interventions in managing GWG by focusing on three key outcomes: total GWG, excessive GWG, and inadequate GWG. This study is a systematic review and meta-analysis to statistically summarize the effect of mobile apps for managing GWG among pregnant women. Furthermore, this study analyzes data from only randomized controlled trials (RCTs) to obtain higher quality evidence. Lastly, by conducting subgroup analyses and meta-regressions, this study aims to investigate the association between the treatment effect of mobile apps and certain variables arising from the trial, participant, intervention characteristics, and control characteristics. This comprehensive investigation into variables that may affect the effectiveness of mobile apps in managing GWG is still lacking and this study aims to bridge this knowledge gap. The findings of this study would be able to guide healthcare providers and inform future efforts on integrating mobile apps adjunct to antenatal care and ultimately improve maternal care in general.

## METHODS

### Search Strategy

The literature search was conducted across six databases, including PubMed, Epistemonikos, EBSCO, Web of Science, Scopus, Google Scholar, and ScienceDirect up to November 9^th^, 2024. The primary keyword searches used in searching for this study were “Application”, “Gestational weight gain”, “Randomized controlled trials”, and their synonyms integrated with boolean operators to expand and specify the search results. The full search strategy in each database can be seen in Appendix 1. No restriction was set for publication year and study language.

### Eligibility Criteria

The studies used refer to the inclusion and exclusion criteria using the Participant, Intervention, Comparison, Outcome, Study Design (PICOS) framework that can be seen in Appendix 2. From this framework, inclusion and exclusion criteria were developed and implemented in screening the studies. Studies to be included fulfil the following requirements: (1) studies that used mobile apps to deliver interventions to promote healthy GWG; (2) randomized controlled trials (RCTs); and (3) RCTs reporting outcomes of absolute GWG from prepregnancy and/or baseline, number of participants that experienced EGWG and/or IGWG during the intervention period. Studies will be excluded if they fulfil one of the following exclusion criterias: (1) full-text is not retrievable; (2) duplicate studies; (3) inappropriate title or abstract to the PICOS framework; (4) intervention being telephone calls or teleconsultation through telephone only; (5) mobile app intervention was started prior to pregnancy; and (6) the article did not report data that can be extracted or used for statistical analysis (this reasoning is hereafter referred to as no extractable data). Grey literature was not excluded and language restriction was employed.

### Study Selection

The collection of study search results from databases was compiled into Rayyan AI (https://www.rayyan.ai/), which has been tested for use in study selection.^7^ Duplication detection was automated and manually screened by removing similar studies with similarity exceeding 90%. The study selection was conducted by two reviewers (GNU, SPN). The “blind-on” mode is activated during the screening process to allow for independent screening of each reviewer to avoid being influenced by other reviewers’ judgement. Once each reviewer completes individual study selection of every study, the “blind-on” mode is deactivated and disagreements are resolved through a discussion between all reviewers (GNU, SPN, FAU). Studies that did not align with the eligibility criteria were removed. The study selection process is reported in the Preferred Reporting Items for Systematic reviews and Meta-Analyses (PRISMA) flow diagram. ^8^

### Data Extraction

Data extraction on studies was conducted to arrange the characteristics of the included studies and perform statistical analysis. The following items were extracted from each study to structure the characteristics of included studies: author, publication year, sample size, participant’s sex, prepregnancy weight, baseline weight, BMI status, prepregnancy BMI, baseline BMI, gestational diabetes mellitus (GDM) status, gestational age, intervention description, control description, and included outcomes for this meta-analysis. The results for each outcome of interest were extracted for statistical analysis.

If studies reported standard error (SE) or 95% confidence interval (CI) instead of standard deviation (SD), the SD was calculated using the formulas recommended by the Cochrane Handbook for Meta-Analysis. If studies reported median and quartiles or range instead of mean and SD, the mean and SD was estimated using the formula proposed by Luo et al.^9^ and Wan et al., 2014^10^, respectively, if there is no significant evidence of non normal distribution of the values tested by the formula proposed by Shi et al.^11^

### Quality Appraisal

Quality assessment of all included studies was done at a study level using Version 2 of the Cochrane risk-of-bias tool for randomized trials (RoB 2), which has five domains and three ratings for studies.^12^ All authors evaluated the study quality separately, and disagreements were resolved between reviewers. The results are compiled and formed with the ROBVIS website as the result of appearance.^13^ Quality assessment was done at the study level.

### Outcomes

This study has two outcomes, which are excessive GWG and inadequate GWG. Excessive GWG and inadequate GWG are defined according to the 2009 Institute of Medicine guideline.^14^

### Statistical Analysis

A random-effect model frequentist meta-analysis was conducted using the “meta” package in R version 4.4.1.^15^ Meta-analysis of excessive GWG and inadequate GWG was conducted using odds ratio (OR). All analysis was performed using a 95% confidence interval (CI). Small-study bias was assessed using Egger’s test and funnel plot visual inspection. Subgroup analysis was conducted by stratifying studies based on categorical data for trial, participant, intervention, and control characteristics. Meta-regression was performed for participant and study characteristics that are continuous variables if they are reported in ≥10 studies.

### Subgroup Analysis and Meta-Regression

Subgroup analyses was planned to compare study-level BMI status inclusion criteria, GDM status inclusion criteria, web versus native apps, social media versus exclusive apps, physical activity targeting interventions versus without, nutrition targeting intervention versus without, self-monitoring features versus without, usual care versus active controls, and continent of the trial.

Meta-regression was planned for prepregnancy weight, baseline weight, prepregnancy BMI, baseline BMI, gestational age at study enrollment, and publication year. Meta-regression was conducted if there are ≥10 studies reporting the mean value of the aforementioned variables.

## RESULTS

### Study Selection

Study identification from the searched databases yielded a total of 1432 articles. After removing 387 duplicates and screening titles/abstracts, 38 potential articles were selected for full-text review. However, the full-text of 3 articles were unable to be retrieved and as such excluded. After a full-text review, 15 RCTs^4,16–29^ were included in this study. The PRISMA flow chart details the study selection process can be seen in Appendix 3.

### Characteristics of Included Studies

A total of 15 RCTs^4,16–29^ published between 2016 and 2024 were included, these studies compare interventions delivered with mobile applications to a standard care group with or without additional non mobile app delivered interventions (control groups with additional interventions are hereby referred to as active control groups). The full study characteristics can be seen in Appendix 4.

### Risk of Bias Assessment

A total of 15 RCTs were quality reviewed using the Cochrane risk-of-bias tool for randomized trials version 2 (RoB 2. The summary plot and traffic light plot containing the risk of bias in each study can be seen in Appendix 5.

### Excessive Gestational Weight Gain

Fifteen RCTs were included for incidence of excessive GWG analysis. Subgroup analysis for physical activity targeting apps and nutrition targeting apps were not able to be performed as all studies included targeted physical activity and nutrition. Only participant age and publication year were reported with mean and SDs in ≥10 RCTs to allow for meta-regression. Meta-analysis showed that mobile apps led to a significant decrease in excessive GWG (OR: 0.71; 95% CI: 0.54 to 0.95; p-value: 0.02; I^2^: 60%; Figure 1). Egger’s regression test found significant small-study bias (p-value: 0.029) and funnel plot visual inspection showed asymmetry (Figure 2).

**Figure 1.**
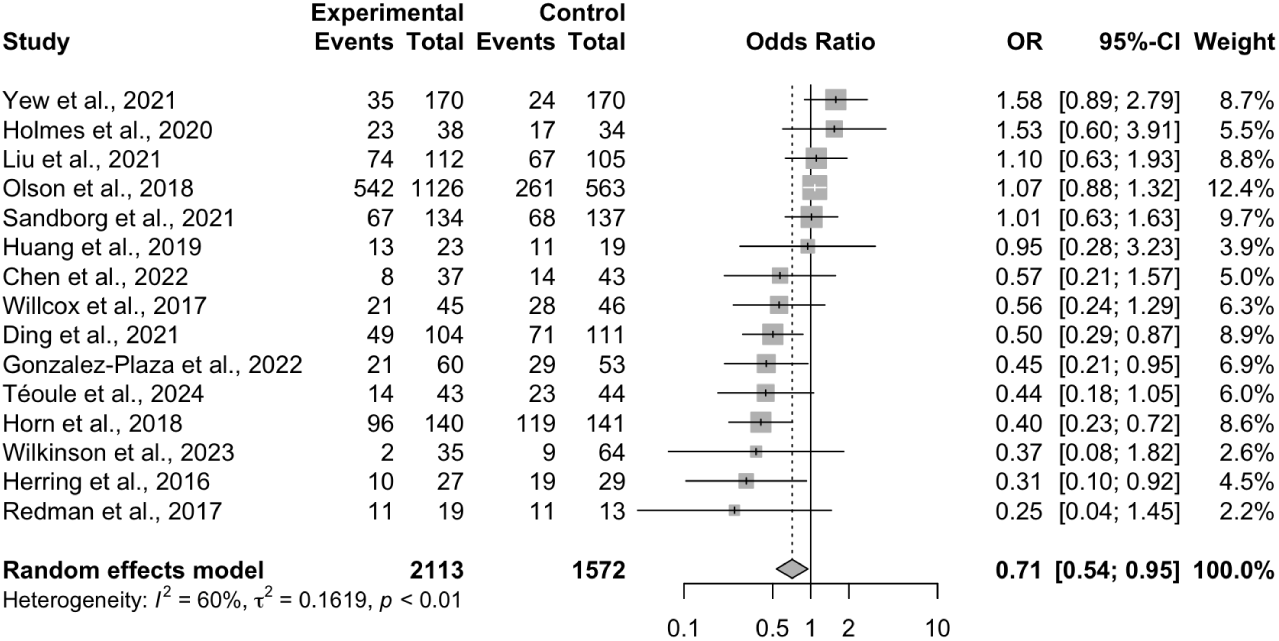
Forest plot for overall analysis of excessive gestational weight gain.

**Figure 2.**
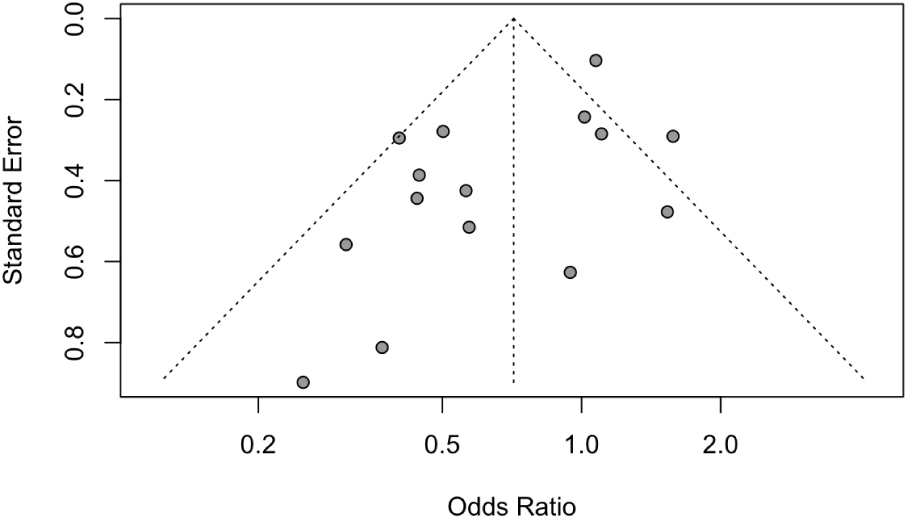
Funnel plot for overall analysis of excessive gestational weight gain.

Subgroup analysis showed that native apps were effective in reducing excessive GWG (OR: 0.63; 95% CI: 0.45 to 0.90; Appendix 6), while web apps were not. Apps were also significant in reducing excessive GWG among studies including exclusively overweight or obese participants (OR: 0.54; 95% CI: 0.33 to 0.90; Appendix 7), while studies that included participants with mixed BMI statuses did not show a significant effect.

When one study with gestational diabetes mellitus (GDM) excluded, the overall efficacy of apps slightly increased (Appendix 8). However, this difference was not statistically significant to the overall analysis as confidence intervals overlap.

Additionally, apps featuring self-monitoring significantly reduced excessive GWG (OR: 0.66; 95% CI: 0.47 to 0.94; Appendix 9), while apps without self-monitoring showed no significant difference. Apps were also effective in reducing excessive GWG when compared exclusively to usual care (OR: 0.65; 95% CI: 0.46 to 0.92; Appendix 10), while studies that compared apps to an active control group showed no significant difference. Subgroup analysis of social media apps versus exclusive apps found that social media was significant in reducing excessive GWG (OR: 0.58; 95% CI: 0.54 to 0.95; Appendix 11), while exclusive apps were not. Subgroup analysis of the trial continent did not reveal significant effects (Appendix 12). Finally, meta-regression indicated that participant age and publication year were not significant moderators of effect size (Appendix 13).

### Inadequate Gestational Weight Gain

Seven RCTs were included for inadequate GWG analysis. Egger’s test, funnel plot visual inspection, and meta-regression were not able to be performed due to inadequate number of studies (<10). Subgroup analysis of GDM status, physical activity targeting apps, nutrition targeting apps, and control group were not able to be performed due to inadequate number of studies in a subgroup.

The overall meta-analysis showed that mobile apps led to a significant increase in inadequate gestational weight gain (OR: 1.51; 95% CI: 1.04 to 2.21; I^2^: 0%; Figure 3).

**Figure 3.**
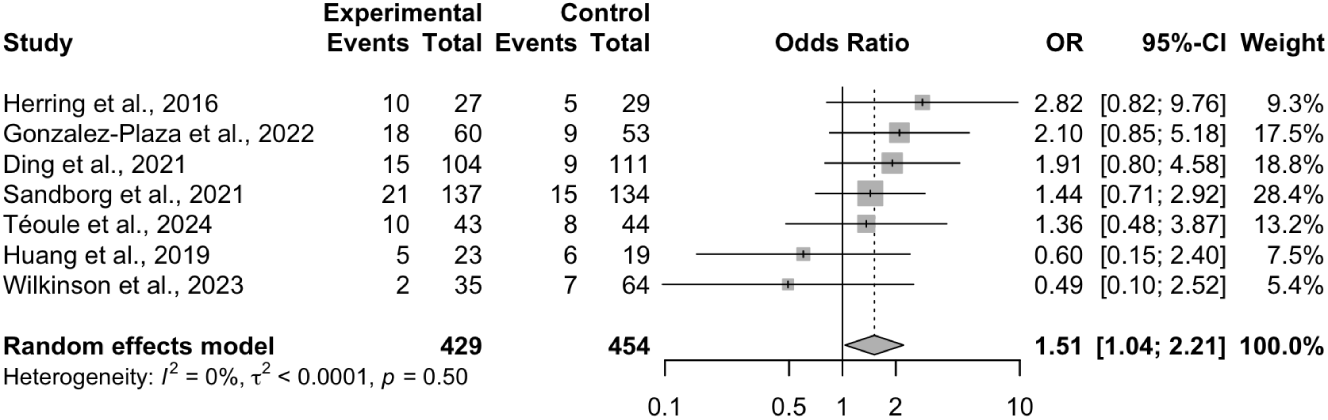
Forest plot for overall analysis of apps on inadequate gestational weight gain.

Subgroup analysis revealed that native apps led to a significantly higher inadequate GWG compared to web apps (OR: 1.94; 95% CI: 1.18 to 3.17; Appendix 14). In contrast, subgroup analyses based on exclusive versus social media apps (Appendix 15), self-monitoring features (Appendix 16), and continent did not show any significant differences (Appendix 17).

## DISCUSSION

To the best of our knowledge, this study is currently the most updated and comprehensive meta-analysis to investigate and summarize the effectiveness of mobile apps in managing gestational weight gain. A previous meta-analysis by Chan et al. found that mobile apps had a significant positive effect on gestational weight management. However, comparing this study to the meta-analysis by Chan et al., which included 8 studies, our meta-analysis incorporated 26 studies, allowing for a more comprehensive evaluation. Furthermore, by separately analyzing total gestational weight gain , dichotomous gestational weight outcomes (excessive and inadequate GWG), and conducting subgroup analyses to explore sources of heterogeneity, our findings provide greater clarity and clinical relevance. This study builds upon what was found in the previous meta-analysis and other studies to develop knowledge on the usage of mobile apps for pregnant women.

The first outcome is the excessive gestational weight gain. The overall analysis revealed mobile app interventions significantly reduced the odds of excessive GWG by 0.71 times. However, this result is limited by significant small-study bias. Thus, there should be high caution in the interpretation of this result and further high quality RCTs are still needed to confirm this finding. Subgroup analyses showed that this significant reduction remained in studies with overweight and obese populations, while vanishing in studies with varied BMI status. This shows that the effectiveness of mobile apps could be higher among overweight and obese patients. Additionally, subgroup analyses found that this significant reduction remained in apps with self-monitoring features, while this effect disappeared in apps that did not. This further puts emphasis that the app’s functionality–particularly self-monitoring–is pivotal. Also, when accounting for the control groups, studies with standard care only had better efficacy. This shows that active control groups may dilute the observed effects and lead to lower treatment effects. Lastly, meta-regression indicated that participant age and publication year were not significant moderators and may not be of concern when considering mobile apps for improving GWG.

The efficacy in social media apps may arise from the ability to give interactive multifaceted interventions (i.e. educational information alongside teleconsultation) at a high intervention dose. Moreover, the capability of social media allows for peer-to-peer support among pregnant women. The effectiveness of self-monitoring functionality may rise from the immediate feedback, self-awareness, and faster monitoring by healthcare providers–which in turn also raises engagement in the intervention–that could further reinforce positive behavioral changes. Furthermore, some studies also incorporated rewards for positive changes as tracked by self-monitoring, which could drive further behavioral change. The underlying reason for the significant result in native apps–however–may not be as clear. It may be due to native apps being more popular than web apps in handheld devices. Even so, it might not even be clear if this effectiveness arises from the native “characteristic” itself. Social media apps–which are shown to be effective–are native apps, thus this effectiveness in native apps may just arise from the effectiveness of social media apps “spilling” over. To ascertain whether native apps are a significant moderator of effect, further studies comparing the web versus native version of the same app are needed.

The second outcome is inadequate gestational weight gain. The overall analysis that mobile app interventions significantly increased the odds of inadequate GWG by 1.51 times. A possible explanation for this finding is the overuse or overemphasis on weight management from the mobile app interventions led to pregnant women excessively constricting their weight gain. This observation is important as it might show that future mobile app interventions may also need to incorporate strategies to avoid undernutrition to combat this risk of inadequate GWG. However, it is also important to note that this result only included seven RCTs. Despite the low heterogeneity (I^2^: 0%) suggesting consistent effects, further studies with larger sample sizes are needed to confirm this finding. Lastly, subgroup analyses indicated that native apps significantly increased the odds of inadequate GWG, while web apps did not. This difference might be due to native apps being significant in reducing total GWG–as mentioned previously in the third paragraph of the discussion–being too excessive and leading to an increase in inadequate GWG.

In essence, mobile app interventions are shown to be effective in reducing excessive GWG. This effectiveness is highlighted by apps that incorporate self-monitoring functionalities. However, the reduction in excessive GWG may be present in only overweight and obese populations, thus further RCTs testing in exclusively normal BMI pregnant women are needed to ascertain this finding. Also, further high quality RCTs are needed to confirm the findings of excessive GWG due to the presence of small-study bias. Lastly, although mobile app interventions are shown to be effective, it comes with an increased risk of inadequate GWG and its implementation should also come with strategies to prevent it.

These findings advances the understanding of the effectiveness of mobile app interventions among pregnant women. The meta-analysis by Chan et al found significant positive effects on birth preparedness knowledge.^6^ The meta-analysis by Wei et al. found significant reduction in fasting blood glucose, postprandial blood glucose, glycated hemoglobin, maternal outcomes, and neonatal outcomes among pregnant women with gestational diabetes mellitus.^30^ Furthermore, an umbrella review by Ameyaw et al. found that mobile apps were also effective in alleviating maternal anxiety and depression, smoking cessation, and controlling substance use during pregnancy.^31^ This study adds reducing excessive GWG to this growing list benefits and acts as further evidence for the support of using mobile app interventions in pregnant women.

This study has several limitations that should be considered when interpreting the results. First, there was high heterogeneity across most analyses and subgroup analyses, likely due to variations in study designs, intervention characteristics, and participant populations. While subgroup analyses and meta-regression were conducted to explore potential sources of heterogeneity, these efforts were insufficient to fully explain the observed variability. Secondly, the small number of included studies and their relatively small sample sizes in the analysis of inadequate GWG limited the statistical power, especially when taking into account the intervention heterogeneity (despite minimal statistical heterogeneity). Third, small-study bias was identified by Egger’s test and funnel plot asymmetry in the analysis of excessive GWG, suggesting that the observed significant reduction in excessive GWG may be overestimated and further larger RCTs are needed to resolve this concern.

There are several knowledge gaps identified by this study. First, the relation between the adherence rate of patients in using the mobile app remains uninvestigated. This study was not able to conduct a meta-regression for adherence rate due to inadequate data. Future RCTs should measure and report adherence rate as well as multivariate analysis incorporating adherence rate as it could be a potential significant moderator of effect. Secondly, long-term post pregnancy outcomes–such as incidence of hypertensive disorders or diabetes mellitus–are still not explored by current studies. Although the reduction of excessive GWG can reduce these long-term complications, it would be better to directly quantify the effectiveness of mobile apps for these complications. Thirdly, due to the characteristic heterogeneity of the interventions, a more standardized intervention would lead to great benefit in synthesizing evidence without heterogeneity concerns.

## CONCLUSION

In conclusion, mobile app interventions are shown to be effective in reducing excessive GWG, particularly social media apps and apps with self-monitoring features. These results suggest that mobile app interventions could be considered for pregnant women to improve maternal and neonatal health. However, the reduction in excessive GWG may only be seen in overweight and obese patients, but more studies are needed to ascertain this finding due to high heterogeneity. LMoreover, further high-quality RCTs are needed to address small-study bias and confirm these effects. Although significant in reducing excessive GWG, their use is associated with an increased risk of inadequate GWG and further modification of the mobile app interventions are needed to combat this risk as it also poses maternal and neonatal complications. Lastly, further studies are needed to investigate the association between adherence rate, long-term post pregnancy outcomes, and develop a more standardized mobile app intervention to reduce heterogeneity.

## Data Availability

All data produced in the present study are available upon reasonable request to the authors

## APPENDIX

**Appendix 1.**
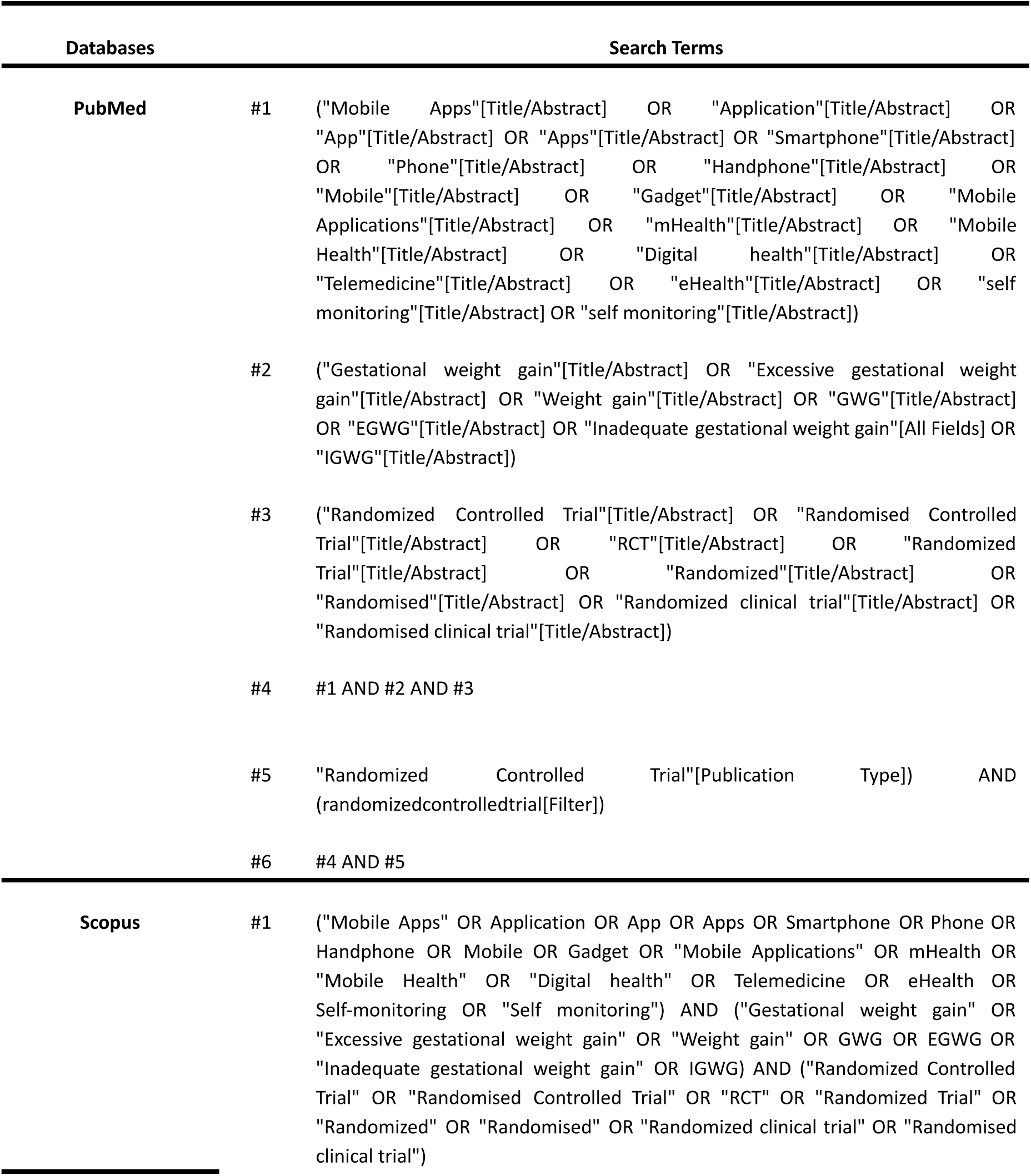
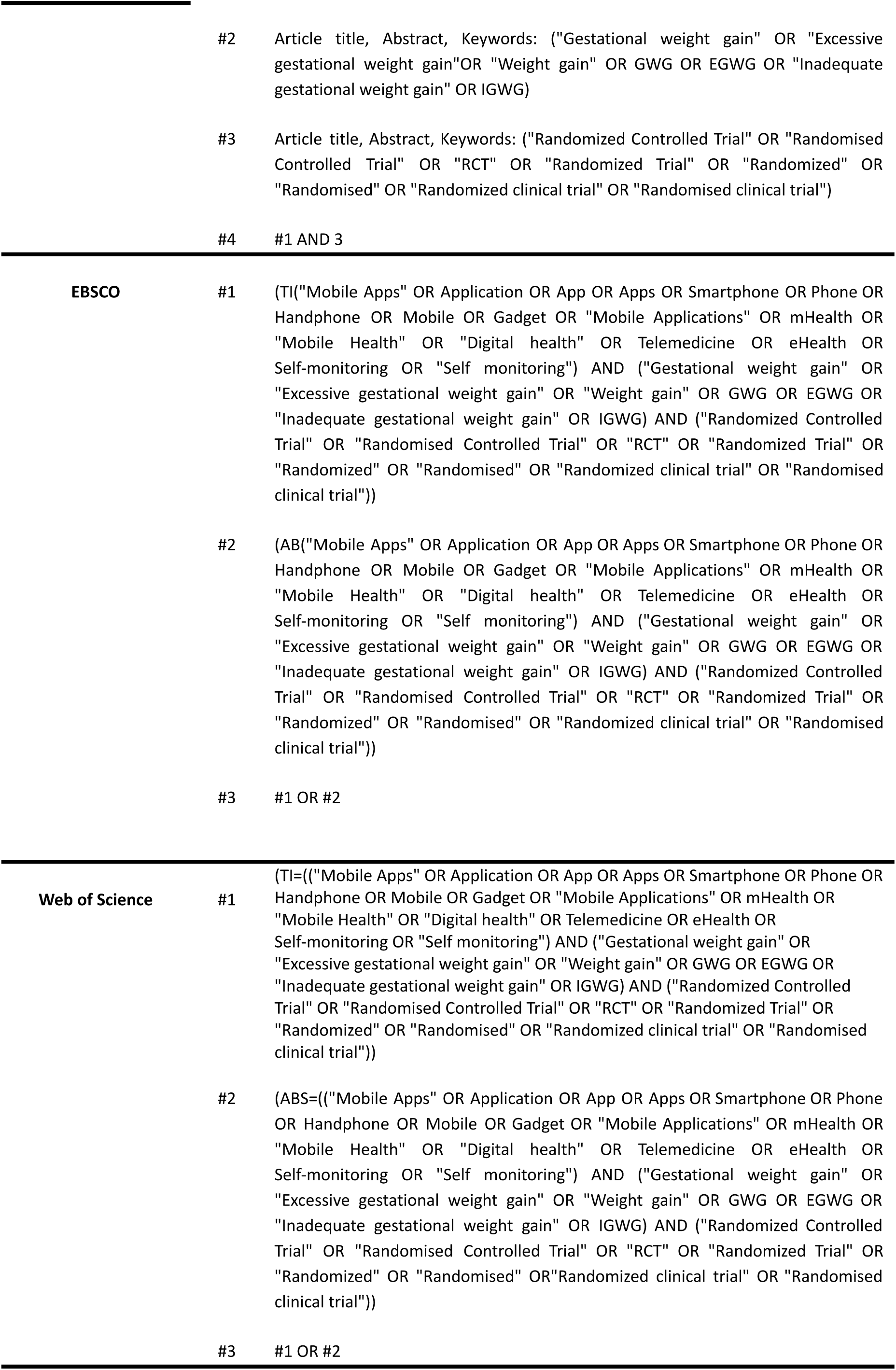
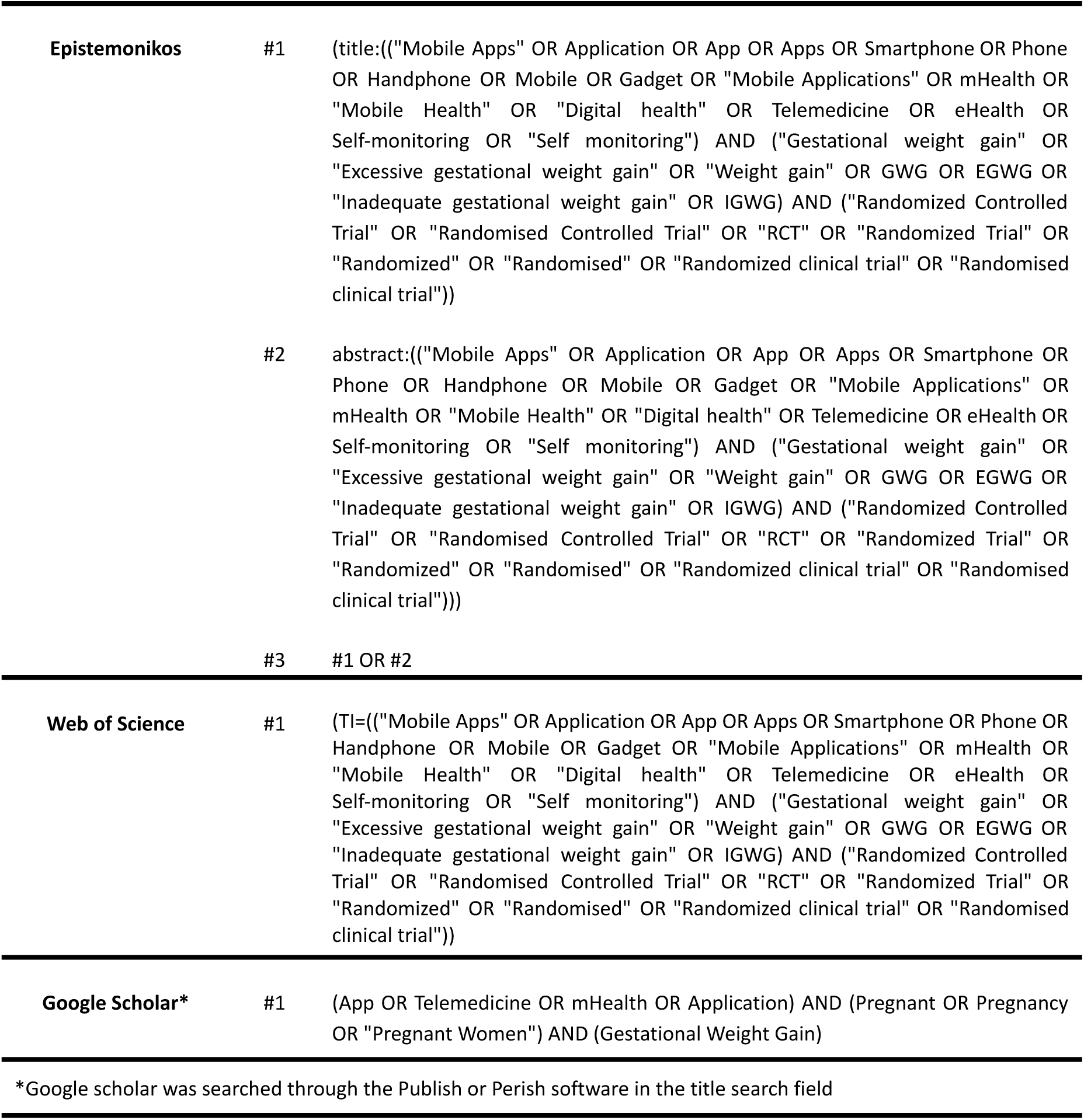
Search strategy used for each database.

**Appendix 2.**
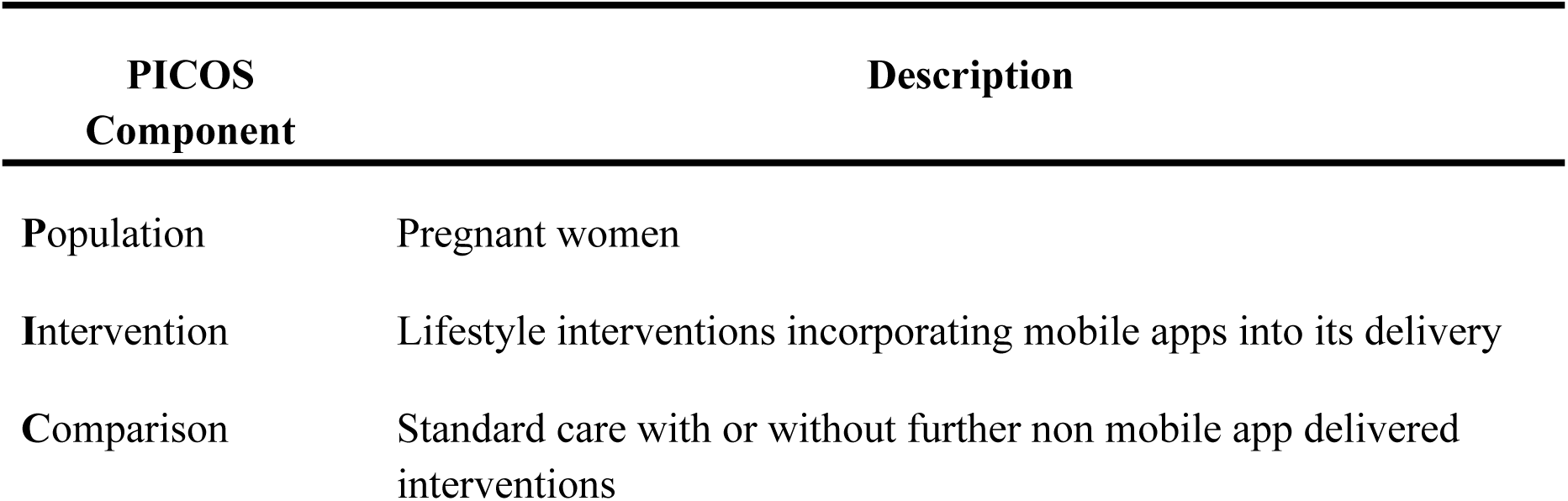
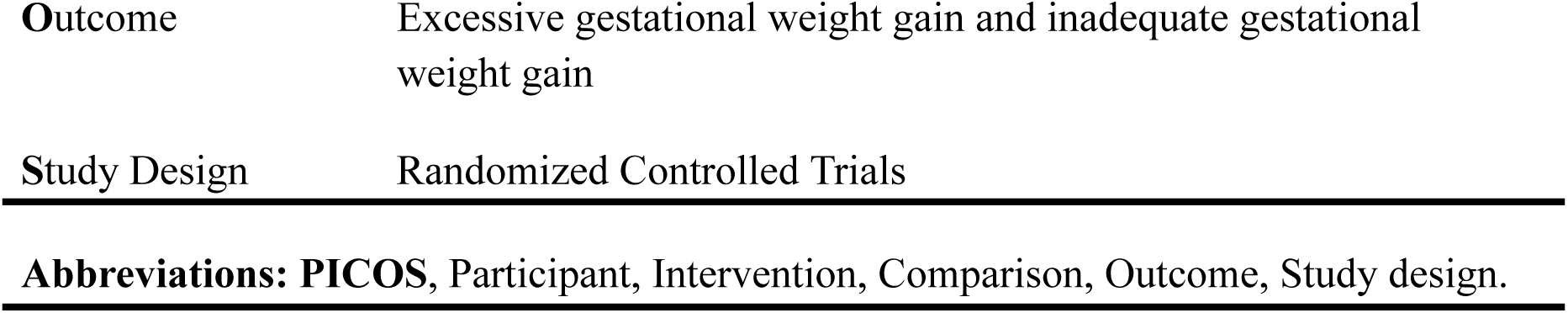
PICOS Framework

**Appendix 3.**
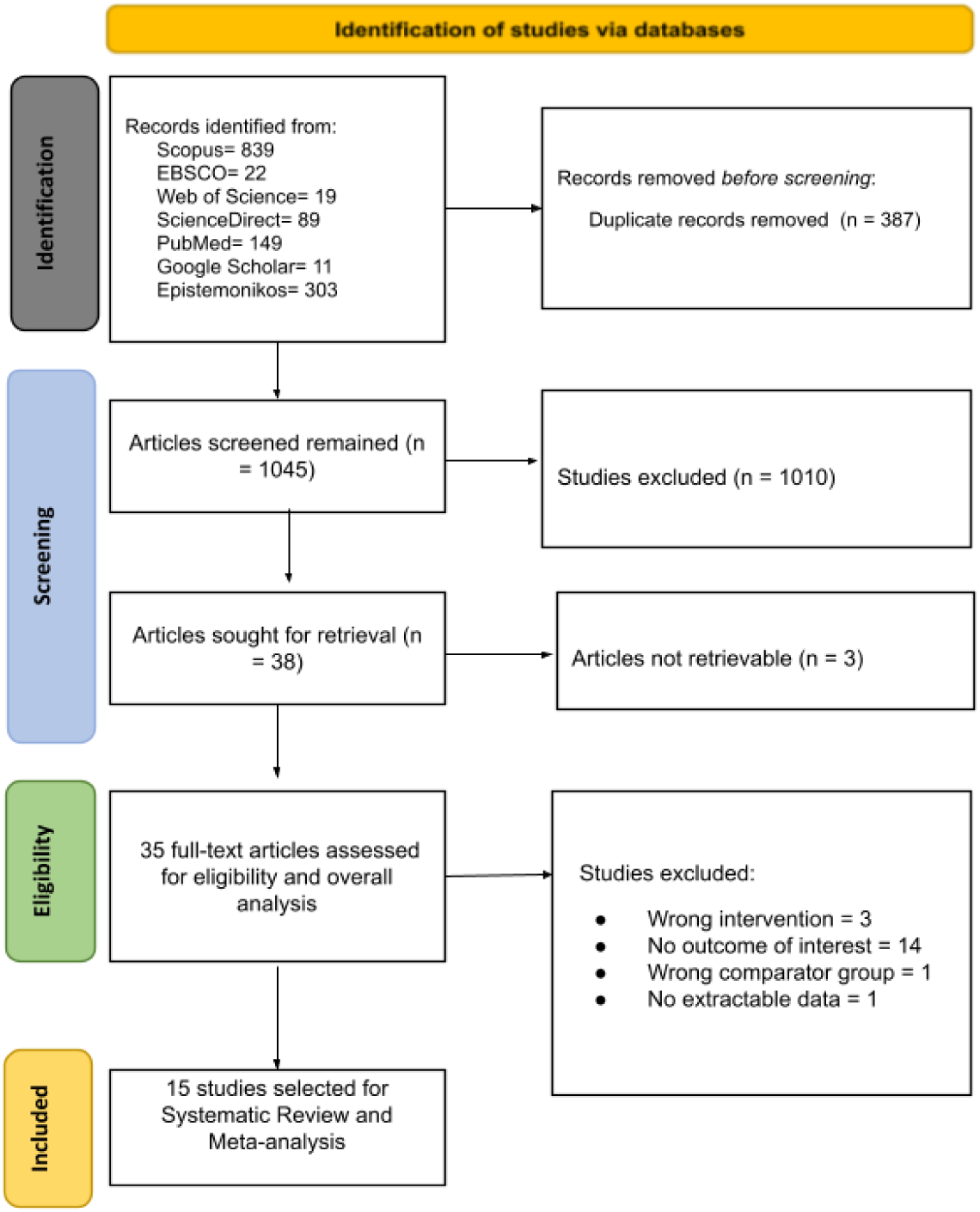
PRISMA flowchart of the study selection

**Appendix 4.** Characteristics of each included study.

| Study | Country | Participant characteristics |  |  |  |  |  |  |  |  | Intervention characteristics |  |  |  | Control Group | Included Outcomes of Interest |
| --- | --- | --- | --- | --- | --- | --- | --- | --- | --- | --- | --- | --- | --- | --- | --- | --- |
|  |  | Sample size | Age | Prepregnancy weight | Baseline Weight | Included BMI status | Baseline BMI | Prepregnancy BMI | GDM status | Baseline Gestational age | Application name | Native or Web | Exclusive or Social Media | Targeted Lifestyle Aspect |  |  |
| Gonzalez-Plaza et al., 2022 | Portugal | 150 | 32.88 ± 5.08 | 85.24 ± 10.17 | NR | Obese | NR | 32.91 ± 3.09 | Any | NR | Mi Fit, Hangouts | Native | Exclusive | Nutrition, Physical Activity, Self-monitoring | Usual care | Excessive GWG, Inadequate GWG |
| Herring et al., 2016 | USA | 66 | 25.45 ± 5.29 | NR | NR | Overweight, Obese | 32.85 ± 5.60 | NR | Any | 12.45 ± 3.65 | Facebook | Native | Social Media | Nutrition, Physical Activity, Self-monitoring | Usual care | Excessive GWG, Inadequate GWG |
| Holmes et al., 2020 | USA | 83 | 27.05 ± 5.42 | 78.42 ± 16.88 | NR | Normal, Overweight, Obese | NR | 30.10 ± 5.71 | Any | NR | EZ Texting | Native | Social Media | Nutrition, Physical Activity | Usual care | Excessive GWG |
| Horn et al., 2018 | USA | 281 | 33.50 ± 4.02 | NR | NR | Overweight, Obese | NR | 31 ± 4 | Any | 13 ± 1 | LOSEIT! | Native | Exclusive | Nutrition, Physical Activity, Self-monitoring | Usual care | Excessive GWG |
| Huang et al., 2019 | Australia | 57 | 33.26 ± 4.16 | NR | 71.69 ± 14.86 | Normal, Overweight, Obese | NR | NR | Any | 9.21 ± 1.21 | PLAN project | Web | Exclusive | Nutrition, Physical Activity, Self-monitoring | Usual care | Excessive GWG, Inadequate GWG |
| Olson et al., 2018 | USA | 1689 | NR | NR | NR | Normal, Overweight, Obese | NR | NR | Any | NR | NR | Web | Exclusive | Nutrition, Physical Activity, Self-monitoring | Usual care + Online health information | Excessive GWG |
| Sandborg et al., 2021 | Sweden | 305 | 31.3 ± 4.1 | NR | 67.7 ± 11.5 | Underweight, Normal, Overweight, Obese | 24.2 ± 3.8 | NR | Any | 13.9 ± 0.7 | HealthyMoms | Native | Exclusive | Nutrition, Physical Activity | Usual care | Excessive GWG, Inadequate GWG |
| Téoule et al., 2024 | Germany | 97 | 32 ± 4 | NR | NR | Normal, Overweight, Obese | NR | 25 ± 4 | Any | NR | Buddy Healthcare & Doctolib | Native | Exclusive | Nutrition, Physical Activity, Self-monitoring, Well-being | Usual care + Wrist fitness tracker | Excessive GWG, Inadequate GWG |
| Wilkinson et al., 2023 | Australia | 99 | 32.70 ± 4.66 | 84.50 ± 15.36 | 85.84 ± 4.86 | Overweight, Obese | NR | 30.68 ± 4.90 | Any | 13.43 ± 3.35 | Propelo | Web | Social Media | Nutrition, Physical Activity, Self-monitoring | Usual care | Excessive GWG, Inadequate GWG |
| Willcox et al., 2017 | Australia | 91 | 32.5 ± 3.4 | NR | NR | Overweight, Obese | NR | 31.0 ± 5.1 | Any | 15.5 ± 2.0 | Facebook | Native | Social Media | Nutrition, Physical Activity, Self-monitoring | Usual care | Excessive GWG |
| Yew et al., 2021 | Singapore | 340 | 32.0 ± 4.2 | NR | NR | Normal, Overweight, Obese | NR | 25.6 ± 5.6 | GDM | 26.9 ± 3.5 | Habits-GDM | Native | Exclusive | Nutrition, Physical Activity, Self-monitoring | Usual care | Excessive GWG |
| Redman et al., 2017 | USA | 36 | 29.24 ± 4.58 | NR | 83.51 ± 12.62 | Overweight, Obese | NR | NR | Any | NR | Fitbit | Native | Exclusive | Nutrition, Physical Activity, Self-monitoring | Usual care | Excessive GWG |
| Liu et al., 2021 | USA | 217 | 29.7 ± 5.0 | NR | 86.9 ± 17.6 | Overweight, Obese | NR | 32.3 ± 5.9 | Any | 12.6 ± 2.3 | MyPlate Daily Checklist for Moms | Web | Exclusive | Nutrition, Physical Activity, Self-monitoring | Usual care + Mailings & Podcasts | Excessive GWG |
| Chen et al., 2022* | Taiwan | 80 | NR | NR | NR | Normal, Overweight, Obese | NR | NR | Any | NR | MHW | Native | Exclusive | Nutrition, Physical Activity, Self-monitoring | Usual care | Excessive GWG |
| Ding et al., 2021 | China | 215 | 30.3 ± 2.8 | 73.8±9.2 | NR | Overweight, Obese | NR | 27.8±2.7 | No GDM | 8.8±1.3 | WeChat | Native | Social Media | Nutrition, Physical Activity, Self-monitoring | Usual care | Excessive GWG, Inadequate GWG |

**Appendix 5.**
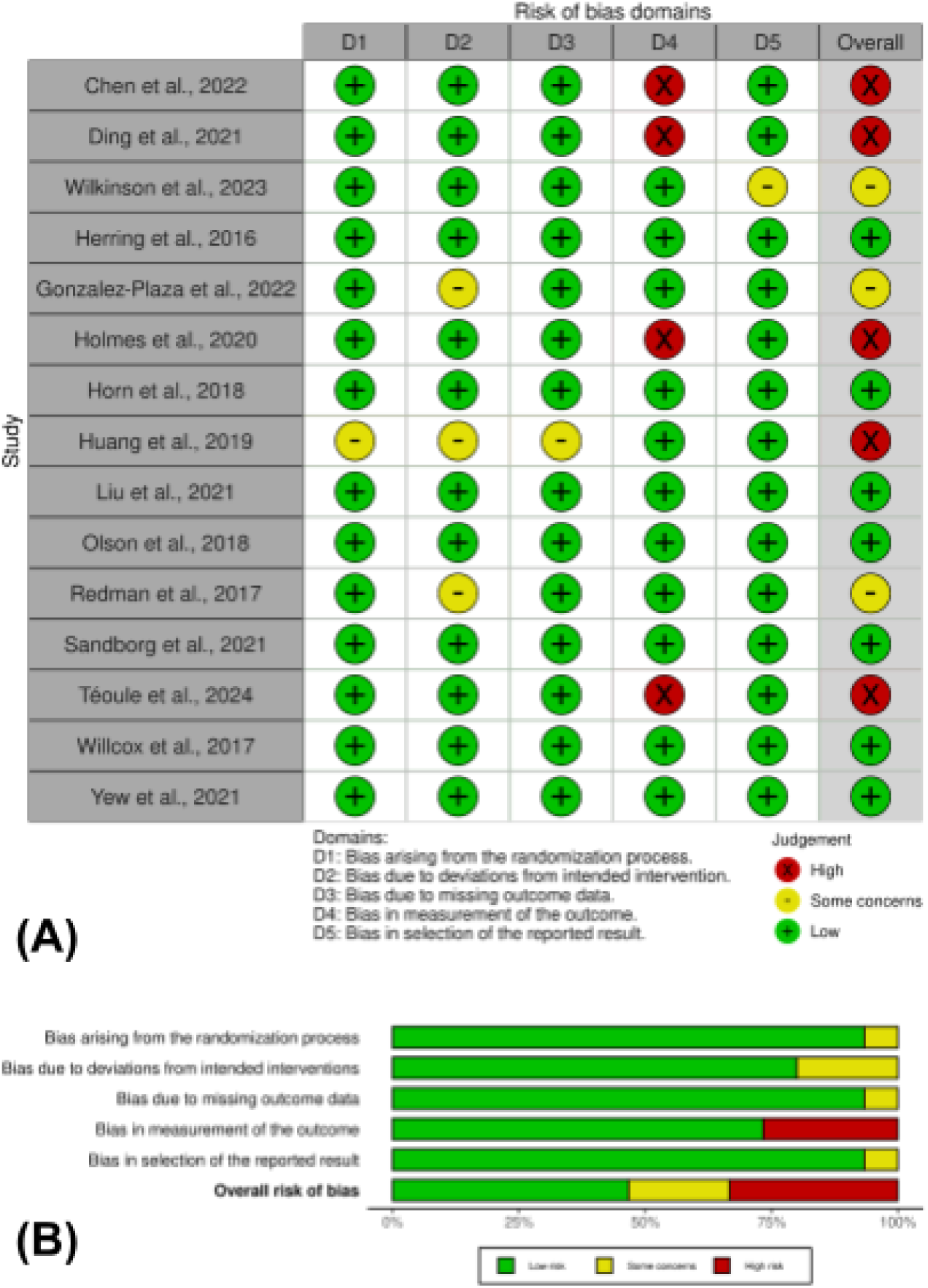
Risk of bias assessment of each included study. (A) Traffic light plot. (B) Summary plot.

**Appendix 6.**
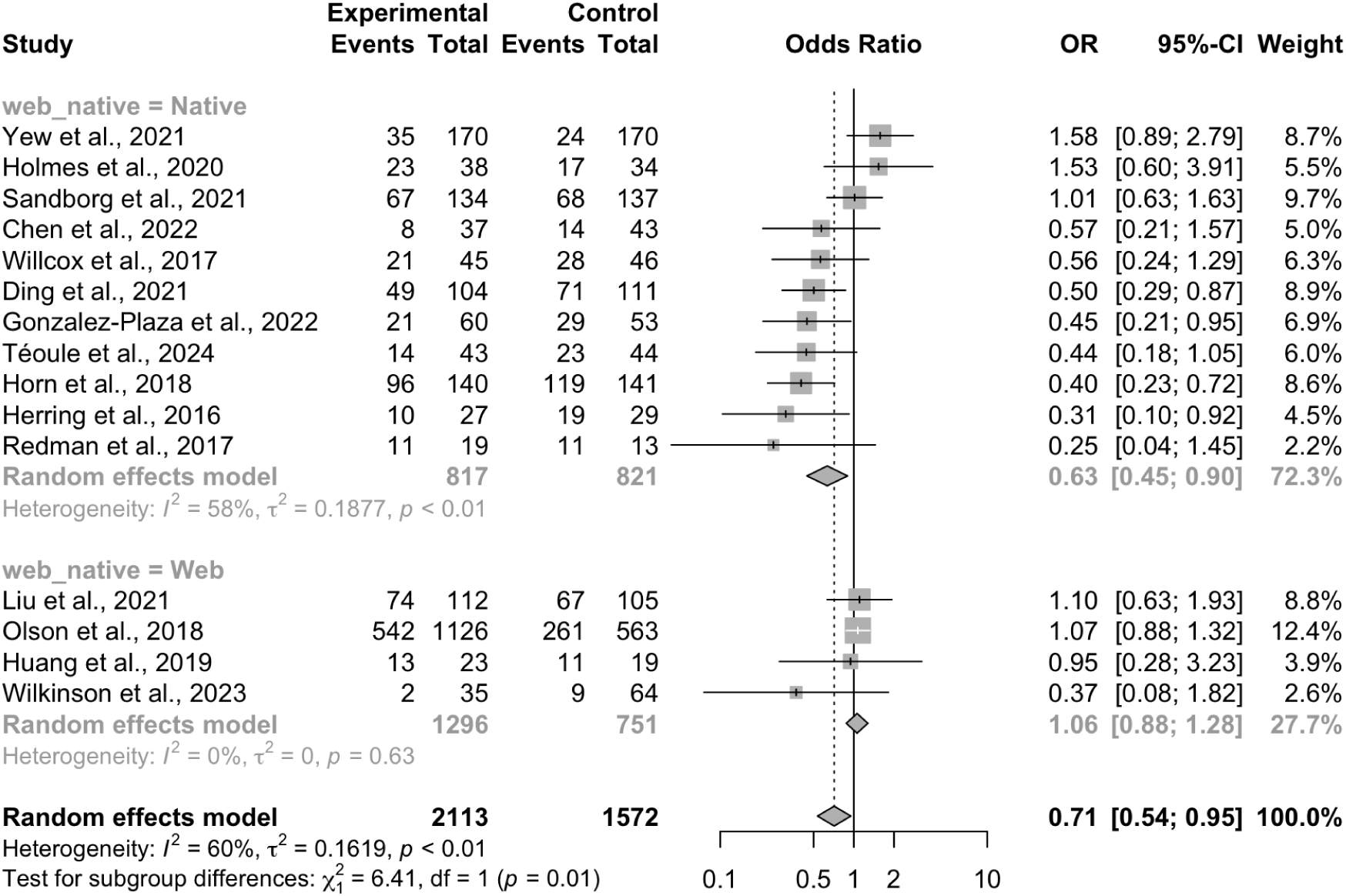
Forest plot for subgroup analysis comparing native to web apps on excessive gestational weight gain.

**Appendix 7.**
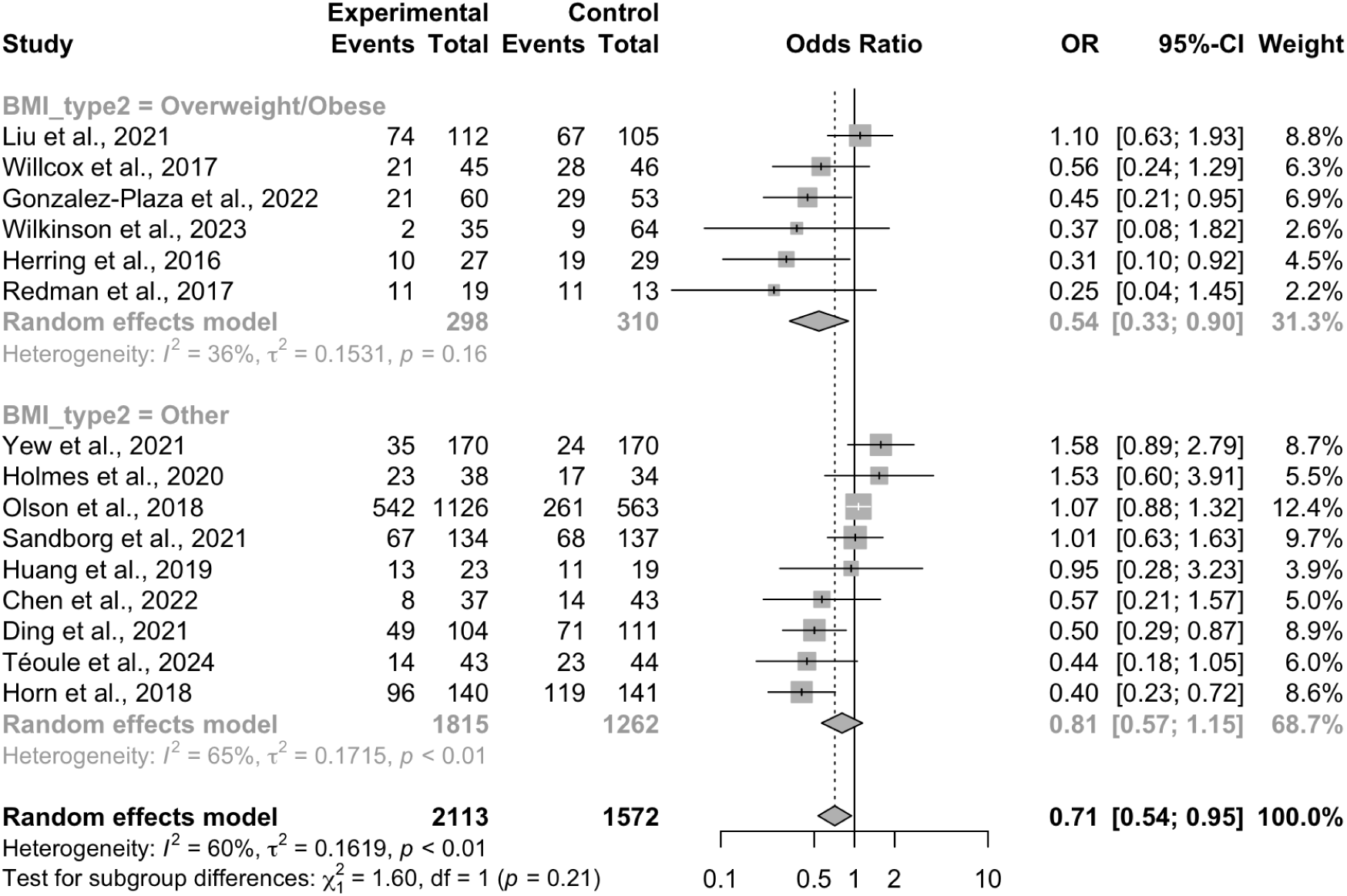
Forest plot for subgroup analysis based on study-level BMI status inclusion criteria on excessive gestational weight gain.

**Appendix 8.**
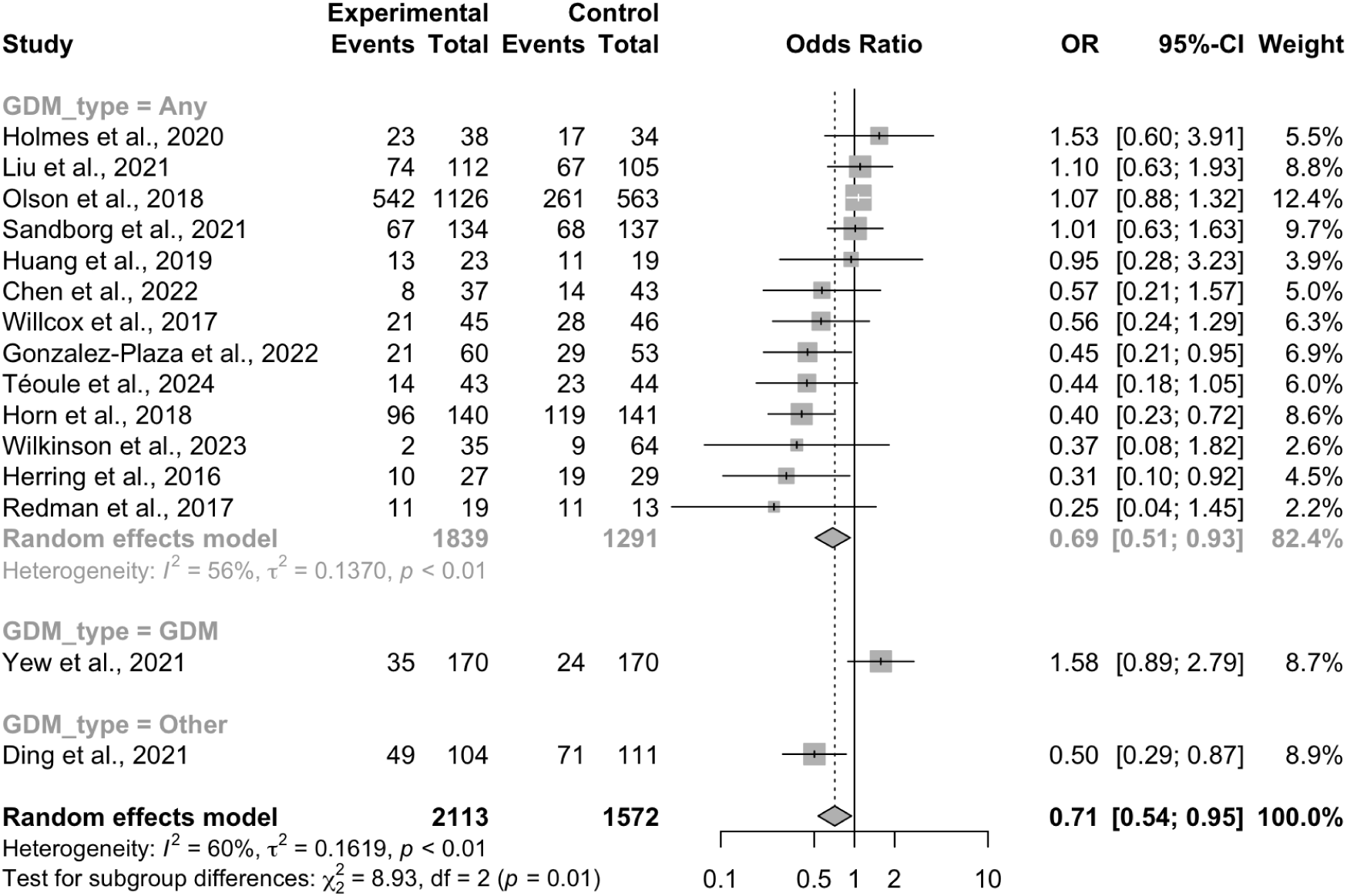
Forest plot for subgroup analysis comparing studies that exclusively included gestational diabetes mellitus patients and those which do not for excessive gestational weight gain. “Other” subgroup refers to studies that excluded pregnant women with gestational diabetes mellitus.

**Appendix 9.**
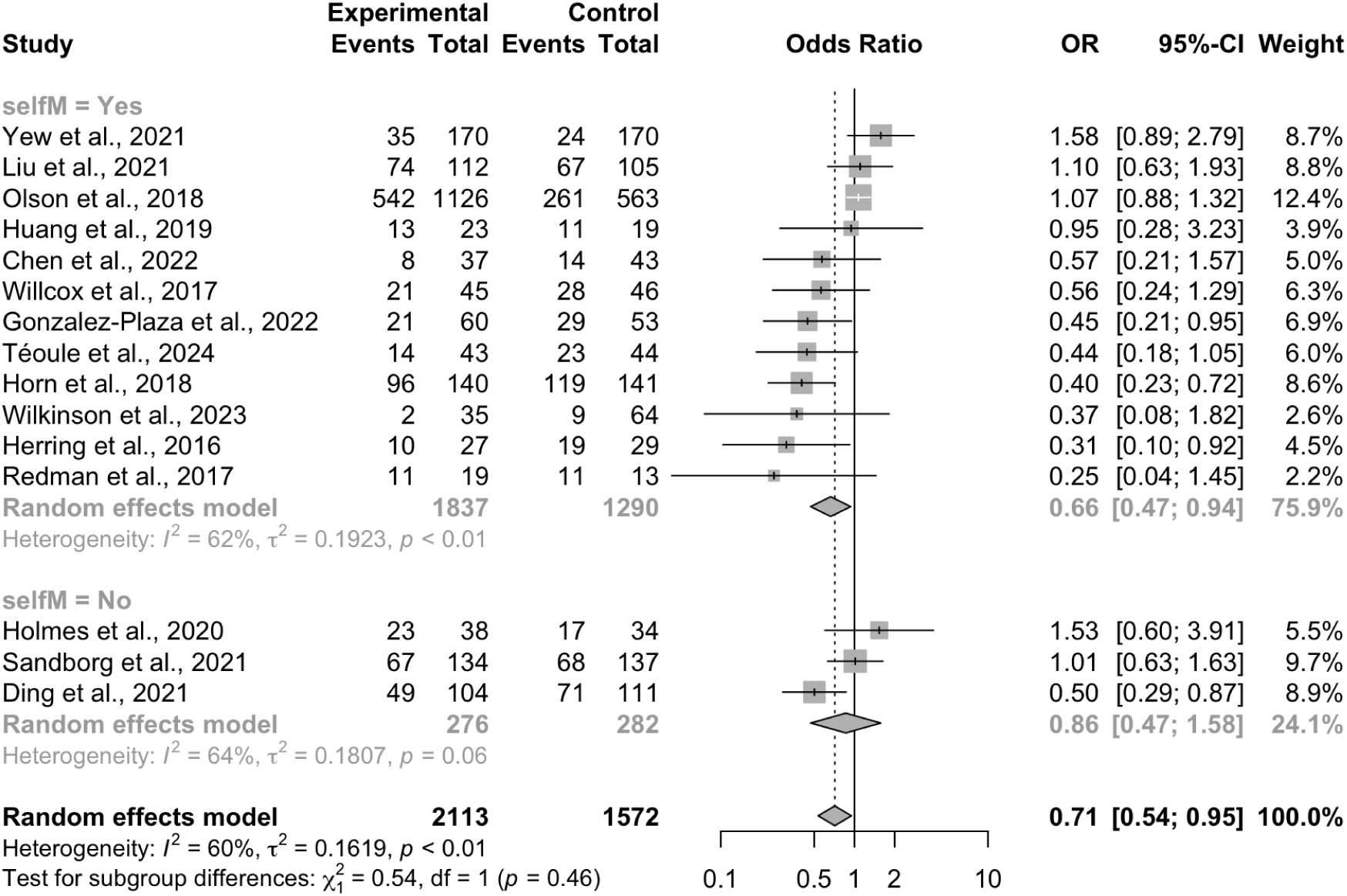
Forest plot for subgroup analysis comparing apps with and without self-monitoring on excessive gestational weight gain.

**Appendix 10.**
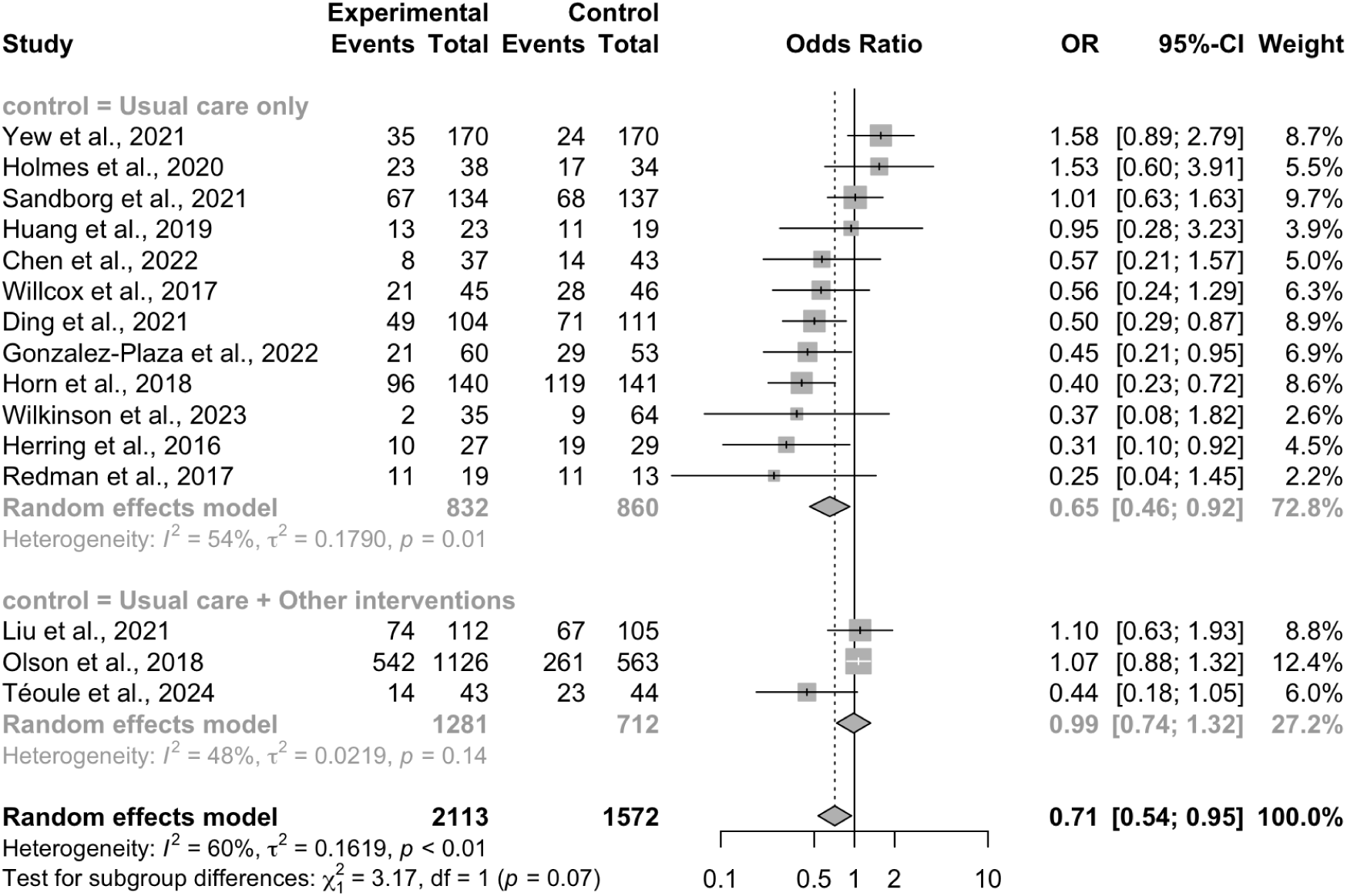
Forest plot for subgroup analysis comparing studies that had standard care only and studies with active control on excessive gestational weight gain.

**Appendix 11.**
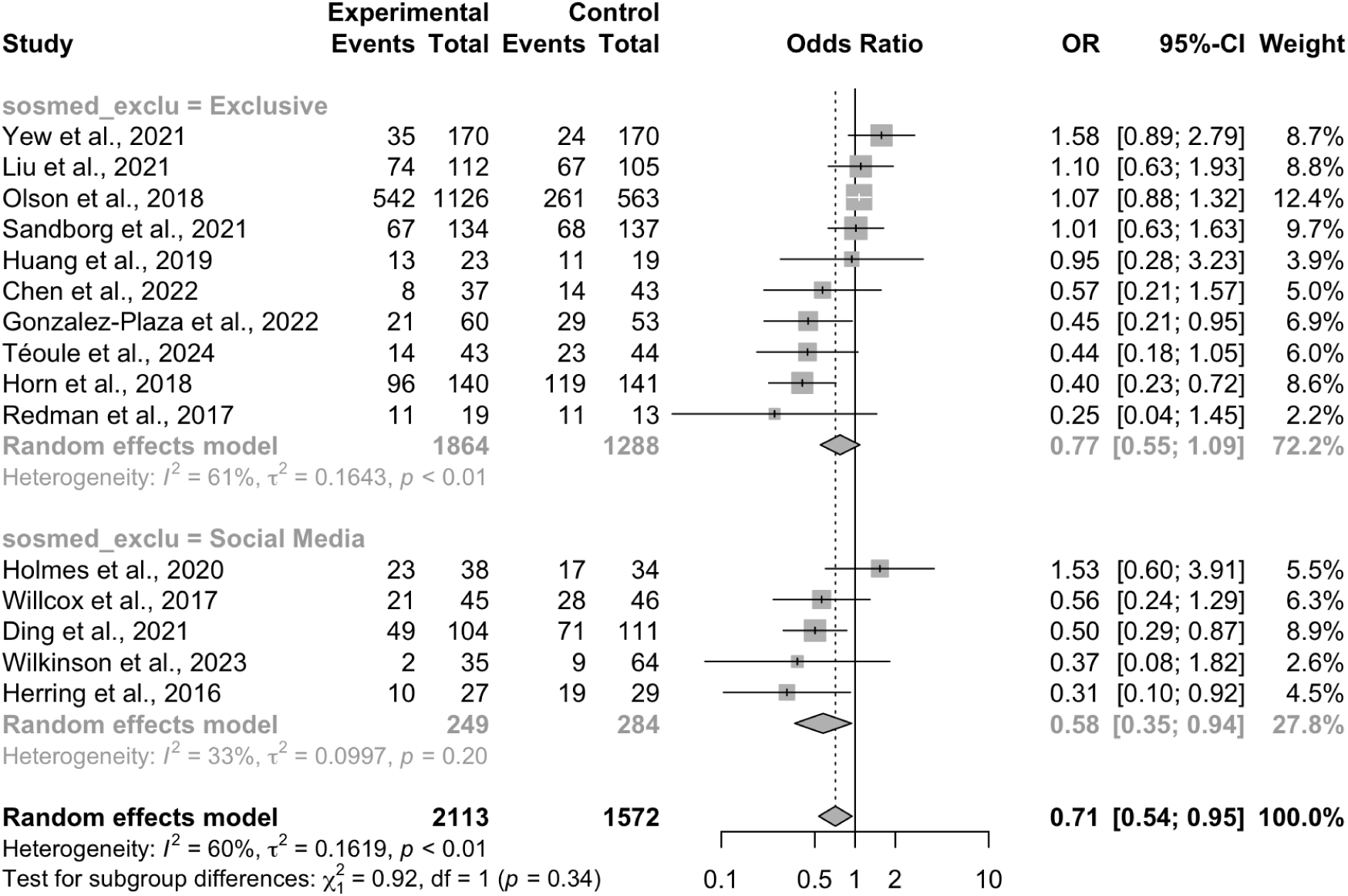
Forest plot for subgroup analysis comparing exclusive apps versus social media apps for excessive gestational weight gain.

**Appendix 12.**
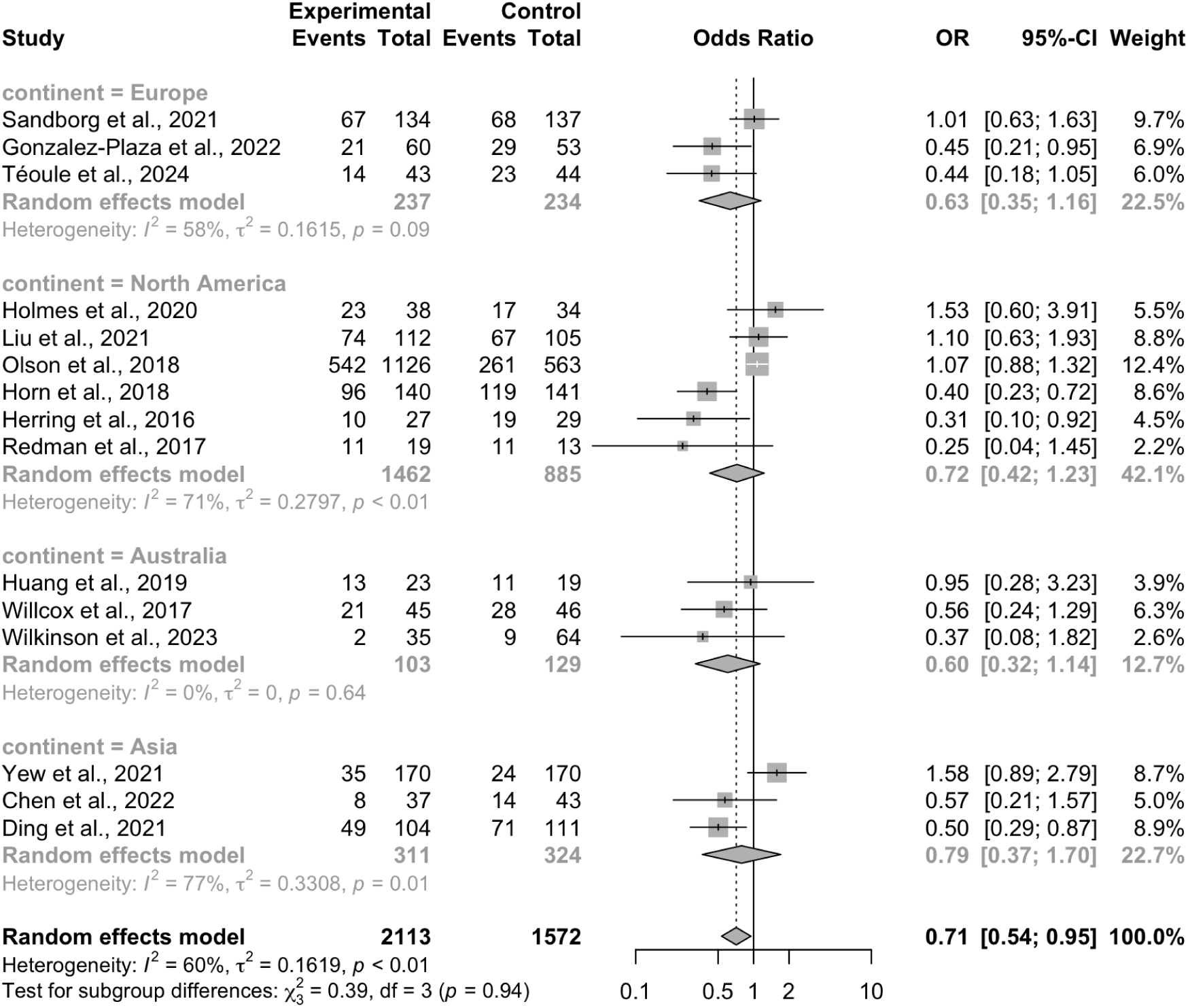
Forest plot for subgroup analysis based on trial continent for excessive gestational weight gain

**Appendix 13.** Results of meta-regression of participant age and publication year for incidence of excessive gestational weight gain. Abbreviation: N, number of studies; SE, standard error; CI, confidence interval

| Moderator | N | Estimate | SE | Lower 95% CI | Upper 95% CI | p-value |
| --- | --- | --- | --- | --- | --- | --- |
| Age | 12 | -0.0226 | 0.0753 | -0.1703 | 0.1251 | 0.7642 |
| Publication year | 14 | 0.0039 | 0.0639 | -0.1213 | 0.1292 | 0.9508 |

**Appendix 14.**
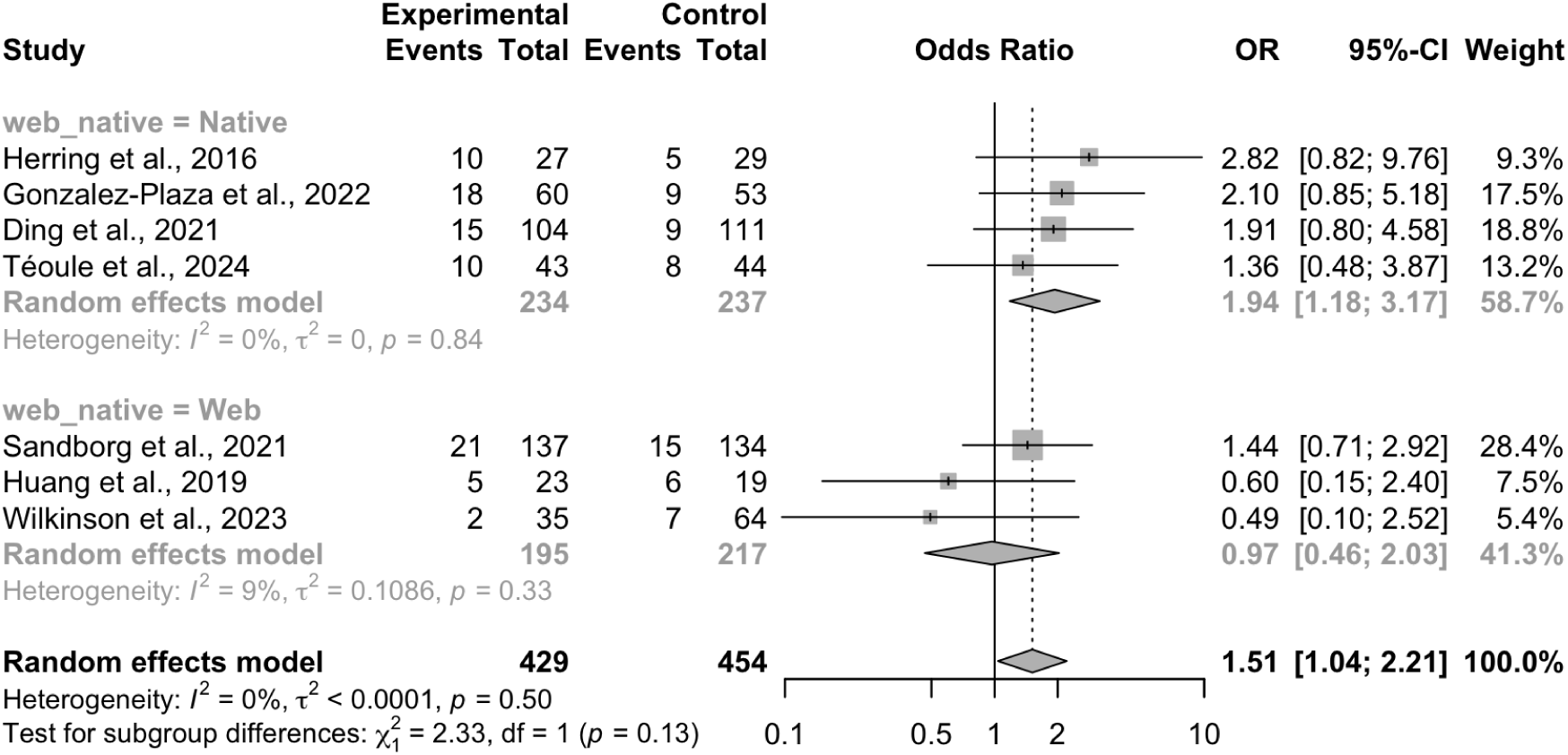
Forest plot for subgroup analysis comparing native apps versus web apps on inadequate gestational weight gain.

**Appendix 15.**
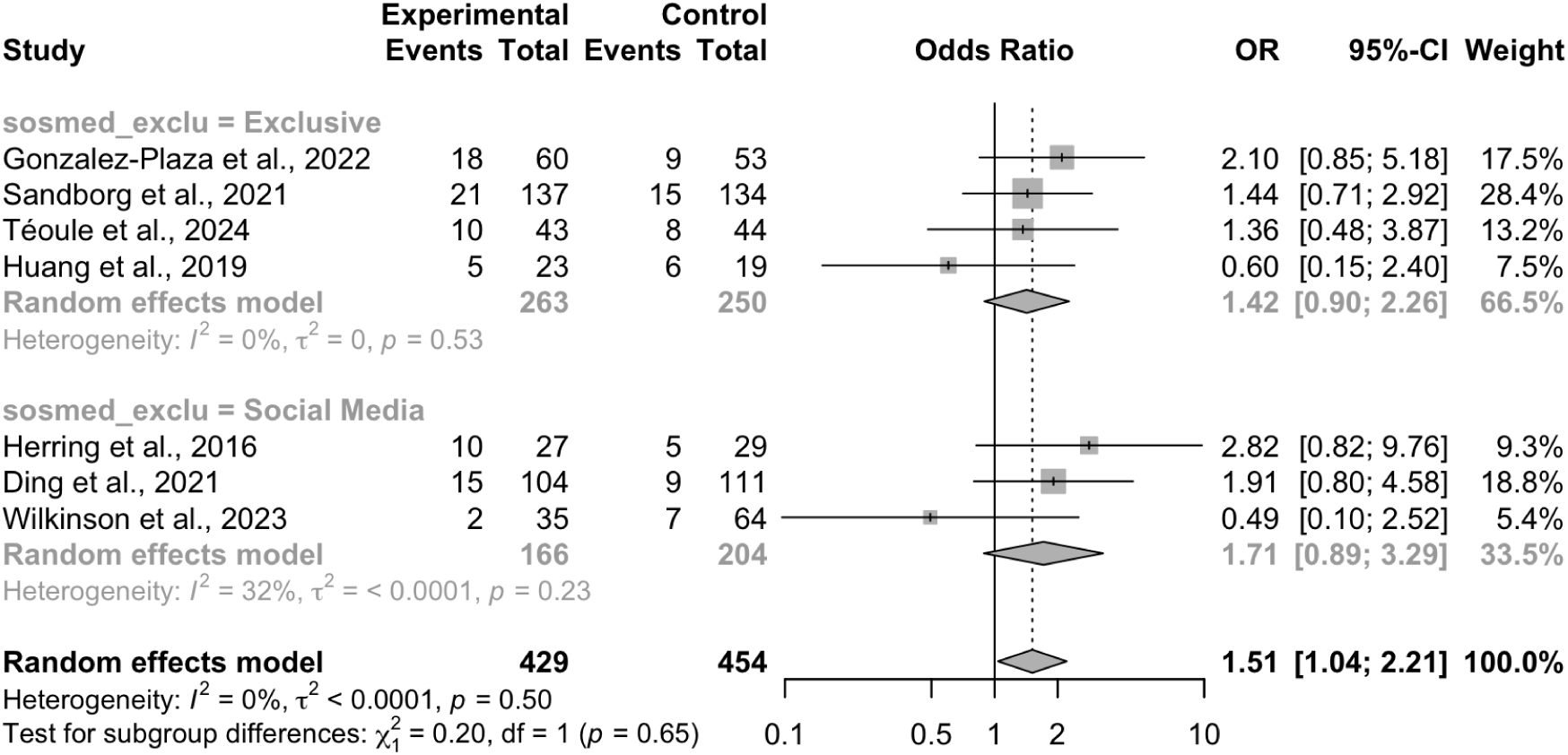
Forest plot for subgroup analysis comparing exclusive apps versus social media apps on inadequate gestational weight gain.

**Appendix 16.**
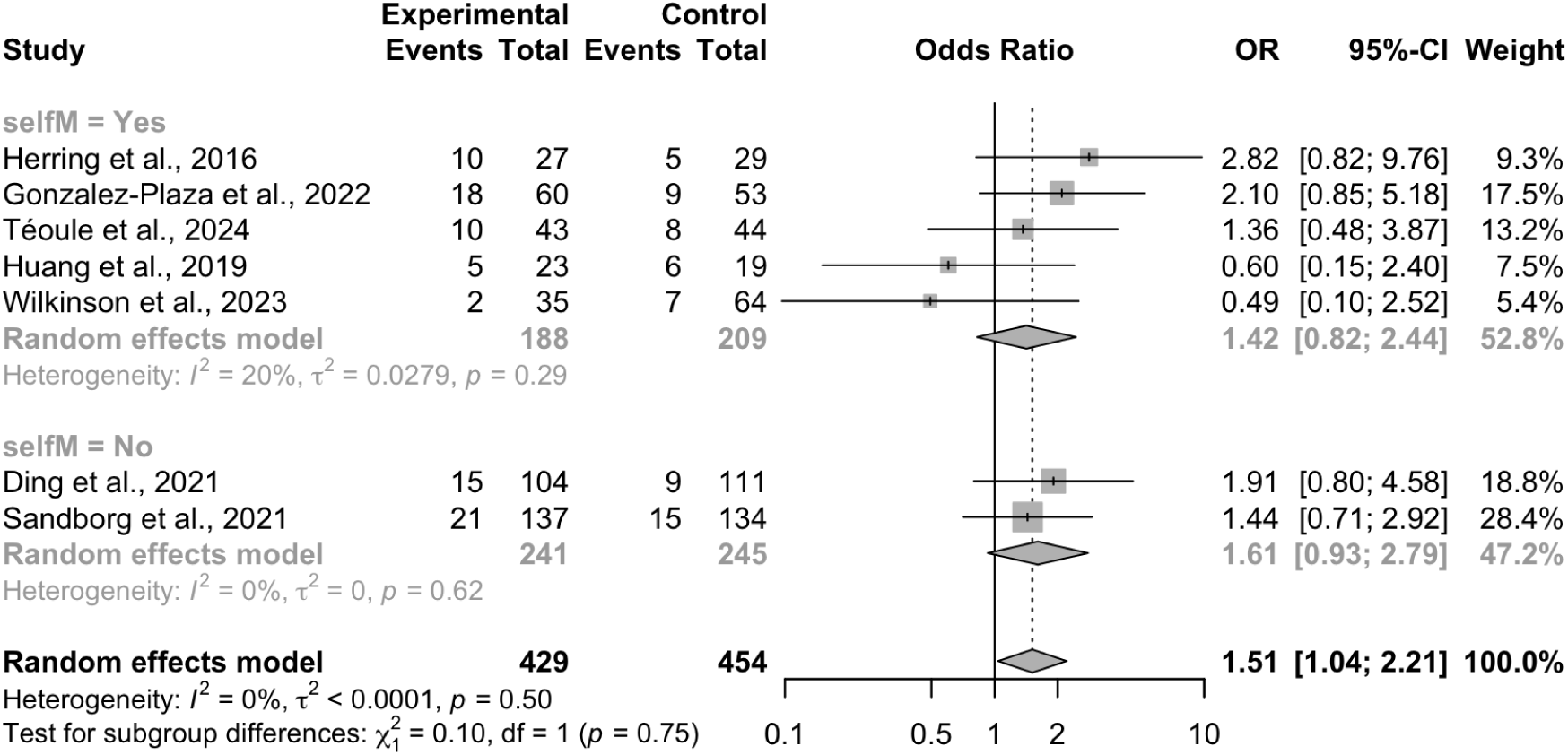
Forest plot for subgroup analysis comparing apps with and without self-monitoring features on inadequate gestational weight gain.

**Appendix 17.**
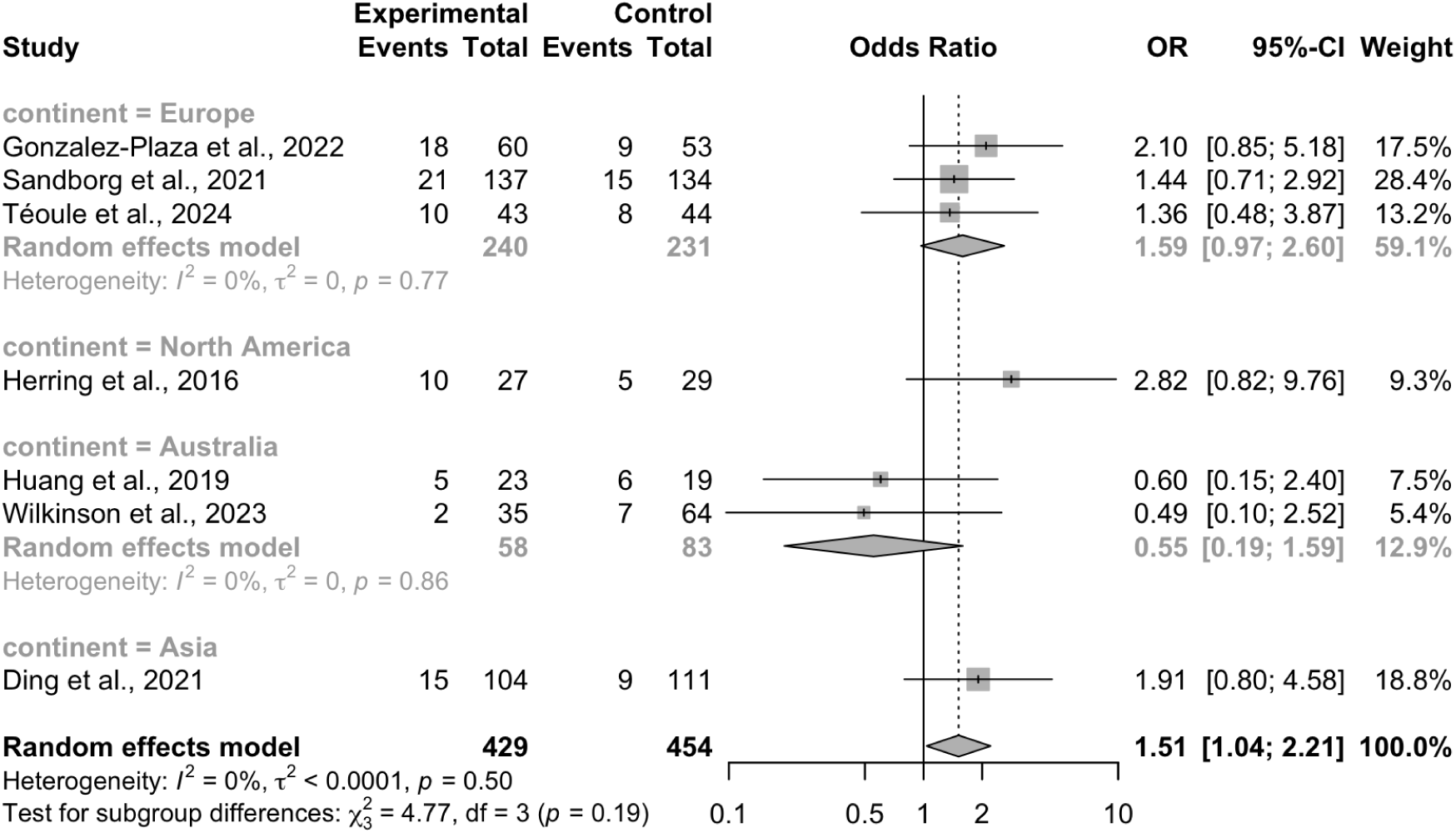
Forest plot for subgroup analysis based on trial continent on inadequate gestational weight gain.

